# Simulated Annealing Identifies Five Shared Drivers of T Cell Exhaustion Across Four Human Cancers

**DOI:** 10.64898/2026.09.15.26363102

**Authors:** Alireza Ebadi, Maryam Hashemi

## Abstract

**Background:** T cell exhaustion is a major barrier to effective cancer immunotherapy. While individual exhaustion markers such as PDCD1 and TOX have been extensively studied, the shared drivers across different cancer types remain poorly defined. Identifying conserved exhaustion drivers could provide pan-cancer therapeutic targets.

**Methods:** We analyzed single-cell RNA-seq data from four human cancers: hepatocellular carcinoma (HCC), colorectal cancer (CRC), melanoma, and non-small cell lung cancer (NSCLC). We applied three complementary computational approaches weighted gene co-expression network analysis (WGCNA), XGBoost-based feature importance, and simulated annealing (SA) — for optimal gene subset selection. Pathway enrichment analysis was performed using KEGG, Reactome, and Gene Ontology (GO) databases.

**Results:** We developed a simulated annealing framework that outperformed WGCNA and XGBoost in identifying conserved exhaustion drivers. SA identified five shared drivers (TOX, PDCD1, HAVCR2, TIGIT, CXCL13) across all four cancers, and twenty novel candidate genes including ITM2A, TNFRSF1B, COTL1, SLA, and PTPN22 that have not been previously linked to exhaustion. Notably, NR4A1 a widely reported exhaustion driver — was not selected in any cancer type, challenging its role as a shared driver. HCC showed a distinct exhaustion signature compared to other cancers.

**Conclusions:** Our study provides a new computational framework (SA) for identifying exhaustion drivers and reveals twenty novel candidate genes. The five shared drivers represent pan-cancer targets, while the novel genes open new avenues for research. The unexpected exclusion of NR4A1 suggests tissue-specific rather than shared roles in exhaustion.

## 1. Introduction

T cell exhaustion is a major barrier to effective cancer immunotherapy, characterized by progressive loss of effector functions, sustained expression of multiple inhibitory receptors, and distinct transcriptional and epigenetic programs. Exhausted CD8+ T cells arise from chronic antigen stimulation in the tumor microenvironment and are regulated by a complex network of transcription factors. This dysfunctional state was first comprehensively defined by Wherry in 2011, who established the conceptual framework for understanding exhaustion as a distinct differentiation state rather than simple anergy or senescence [1]. Subsequent work by Wherry and Kurachi in 2015 provided a detailed molecular and cellular characterization of exhausted T cells, identifying key transcriptional regulators and surface markers that distinguish them from effector and memory T cells [2]. Blank and colleagues further refined the definition of T cell exhaustion in 2019, proposing a consensus framework that distinguishes exhaustion from other forms of T cell dysfunction such as anergy and senescence [3]. The clinical significance of T cell exhaustion has been extensively documented. McLane and colleagues demonstrated that CD8+ T cell exhaustion occurs during chronic viral infection and cancer, with exhausted cells showing progressive loss of proliferative capacity and effector function [4]. Pauken and Wherry showed that overcoming T cell exhaustion is critical for effective immunotherapy, as exhausted cells fail to respond to checkpoint blockade [5]. Hashimoto and colleagues further demonstrated that CD8+ T cell exhaustion represents a major barrier to cancer immunotherapy, with exhausted cells showing distinct metabolic and transcriptional profiles [6]. Thommen and Schumacher provided a comprehensive overview of T cell dysfunction in cancer, highlighting the role of inhibitory receptors and metabolic constraints [7]. The inhibitory receptors that define exhaustion have been extensively characterized. Anderson and colleagues demonstrated that LAG-3, TIM-3, and TIGIT are co-inhibitory receptors with specialized functions in immune regulation, each contributing to distinct aspects of T cell dysfunction [8]. Barber and colleagues showed that PD-1 blockade can restore function in exhausted CD8+ T cells during chronic viral infection, providing the first evidence that exhaustion is reversible [9]. Day and colleagues demonstrated that PD-1 expression on HIV-specific T cells is associated with T cell exhaustion and disease progression, establishing PD-1 as a key marker of exhaustion [10]. Wherry and colleagues further defined the molecular signature of CD8+ T cell exhaustion during chronic viral infection, identifying a distinct transcriptional program that includes multiple inhibitory receptors [11]. Blackburn and colleagues showed that co-regulation of CD8+ T cell exhaustion by multiple inhibitory receptors occurs during chronic viral infection, with PD-1 and LAG-3 acting synergistically [12]. The role of specific inhibitory receptors in cancer has been extensively studied. Fourcade and colleagues demonstrated that upregulation of TIM-3 and PD-1 is associated with tumor antigen-specific CD8+ T cell dysfunction in melanoma patients [13]. Spranger and colleagues showed that upregulation of PD-L1, IDO, and Tregs in the melanoma microenvironment is driven by CD8+ T cells, establishing a feedback loop that sustains exhaustion [14]. Pauken and colleagues demonstrated that the epigenetic stability of exhausted T cells limits the durability of reinvigoration by PD-1 blockade, showing that exhaustion is maintained by stable epigenetic modifications [15]. Sen and colleagues further characterized the epigenetic landscape of T cell exhaustion, identifying specific chromatin states that define exhausted cells [16]. Philip and colleagues showed that chromatin states define tumor-specific T cell dysfunction and reprogramming, providing a framework for understanding how exhaustion is established and maintained [17]. The transcription factor TOX has emerged as a central regulator of exhaustion. Khan and colleagues demonstrated that TOX transcriptionally and epigenetically programs CD8+ T cell exhaustion, identifying TOX as a master regulator that recruits chromatin remodelers to maintain exhaustion-associated gene expression [18]. Alfei and colleagues showed that TOX reinforces the phenotype and longevity of exhausted T cells in chronic viral infection, demonstrating that TOX is required for the persistence of exhausted cells [19]. Scott and colleagues demonstrated that TOX is a critical regulator of tumor-specific T cell differentiation, showing that TOX expression increases during tumor progression [20]. Yao and colleagues used single-cell RNA-seq to reveal TOX as a key regulator of CD8+ T cell persistence in chronic infection [21]. Seo and colleagues demonstrated that TOX and TOX2 cooperate with NR4A transcription factors to impose CD8+ T cell exhaustion, identifying a transcriptional network that drives exhaustion [22]. The NR4A family of transcription factors has also been implicated in exhaustion. Liu and colleagues performed a genome-wide analysis that identified NR4A1 as a key mediator of T cell dysfunction, demonstrating that NR4A1 regulates genes involved in exhaustion [23]. Chen and colleagues showed that NR4A transcription factors limit CAR T cell function in solid tumors, providing evidence that NR4A family members restrict antitumor immunity [24]. Martínez and colleagues demonstrated that the transcription factor NFAT promotes exhaustion of activated CD8+ T cells, identifying NFAT as an upstream regulator of the exhaustion program [25]. These studies collectively establish NR4A1 as a critical mediator of T cell dysfunction, making its exclusion from our shared drivers particularly noteworthy. Single-cell RNA sequencing has transformed our understanding of T cell states in cancer. Thommen and colleagues identified a transcriptionally and functionally distinct PD-1+ CD8+ T cell pool with predictive potential in non-small-cell lung cancer treated with PD-1 blockade [26]. Guo and colleagues provided a global characterization of T cells in non-small-cell lung cancer by single-cell sequencing, identifying distinct T cell subsets with different functional states [27]. Sade-Feldman and colleagues defined T cell states associated with response to checkpoint immunotherapy in melanoma, identifying distinct exhausted T cell populations [28]. Li and colleagues showed that dysfunctional CD8 T cells form a proliferative, dynamically regulated compartment within human tumors, revealing the heterogeneity of exhausted T cells [29]. Miller and colleagues demonstrated that subsets of exhausted CD8+ T cells differentially mediate tumor control and respond to checkpoint blockade, identifying progenitor and terminal exhausted populations [30]. The spatial organization of exhausted T cells has emerged as a critical determinant of their function. Jansen and colleagues demonstrated that an intra-tumoral niche maintains and differentiates exhausted T cell states, showing that exhausted T cells are not randomly distributed [31]. Siddiqui and colleagues showed that intratumoral Tcf1+PD-1+CD8+ T cells with stem-like properties promote tumor control in response to vaccination and checkpoint blockade [32]. Im and colleagues defined the CD8+ T cells that provide the proliferative burst after PD-1 therapy, identifying a stem-like population that gives rise to exhausted cells [33]. Huang and colleagues demonstrated that the ratio of T-cell invigoration to tumour burden is associated with anti-PD-1 response, providing a clinical correlate of exhausted T cell dynamics [34]. van der Leun and colleagues provided a comprehensive review of CD8+ T cell states in human cancer, integrating single-cell analysis with clinical outcomes [35]. The tumor microenvironment plays a central role in shaping T cell exhaustion. Zhang and colleagues demonstrated that TGF-β signaling in the tumor microenvironment contributes to T cell exhaustion, with TGF-β suppressing effector function and promoting exhaustion [36]. Liu and colleagues used temporal single-cell tracing to reveal clonal revival and expansion of precursor exhausted T cells during anti-PD-1 therapy in lung cancer [37]. Song and colleagues performed single-cell meta-analyses that revealed responses of tumor-reactive CXCL13+ T cells to immune-checkpoint blockade, identifying CXCL13 as a key marker of exhausted T cells [38]. Kim and colleagues provided a comprehensive single-cell map of T cell exhaustion-associated immune environments in human breast cancer [39]. The tumor microenvironment of specific cancers has been characterized by single-cell sequencing. Zheng and colleagues provided a landscape of infiltrating T cells in liver cancer revealed by single-cell sequencing, identifying distinct T cell states in hepatocellular carcinoma [40]. Zhang and colleagues characterized the landscape and dynamics of single immune cells in hepatocellular carcinoma, revealing the heterogeneity of immune cells in this cancer type [41]. Kim and colleagues used single-cell RNA sequencing to demonstrate the molecular and cellular reprogramming of the tumor microenvironment in human colorectal cancer [42]. These studies establish the context-specific nature of T cell exhaustion in different cancer types. The current study builds on this extensive body of work. Blank and Haining provided a conceptual framework for defining T cell exhaustion, which we used to guide our analysis [43]. Pauken and Wherry discussed the role of T cell exhaustion in chronic viral infection and cancer, providing a foundation for understanding the clinical implications of our findings [44]. Wherry and Kurachi provided a comprehensive review of the molecular and cellular insights into T cell exhaustion, which informed our selection of exhaustion markers [45]. The study by Khan and colleagues established TOX as a central regulator of exhaustion, providing a benchmark for our computational framework [46]. The work by Alfei and colleagues further demonstrated the importance of TOX in maintaining exhausted T cells [47]. The study by Scott and colleagues characterized TOX as a critical regulator of tumor-specific T cell differentiation, which we used to validate our findings [48]. The work by Seo and colleagues identified the TOX-NR4A transcriptional network, which we examined in our pan-cancer analysis [49]. Finally, the study by Liu and colleagues identified NR4A1 as a key mediator of T cell dysfunction, making its exclusion from our shared drivers particularly significant [50].

## 2. Methods

### 2.1 Data collection

Single-cell RNA-seq data from four human cancers were obtained from public repositories. The datasets included:

| Cancer | Dataset ID | Source | Cells | Genes | Platform |
| --- | --- | --- | --- | --- | --- |
| HCC | GSE98638 | GEO | 5,064 | 23,459 | Illumina HiSeq 2500 |
| CRC | GSE108989 | GEO | 11,139 | 23,689 | Illumina HiSeq 4000 |
| Melanoma | GSE72056 | GEO | 4,645 | 23,689 | Illumina HiSeq 2500 |
| NSCLC | GSE99254 | GEO | 12,346 | 23,459 | Illumina HiSeq 4000 |

All datasets were downloaded from the NCBI Gene Expression Omnibus (GEO) database. Raw count matrices were provided as txt.gz files. For each dataset, the count matrix was parsed using pandas with chunked reading to manage memory. Gene symbols were extracted from the symbol column and used as feature names. The expression matrix was transposed to obtain a cells × genes format.

### 2.2 Quality control and preprocessing

Cells were filtered to retain those with at least 200 detected genes and less than 20% mitochondrial reads. Genes expressed in fewer than 3 cells were excluded. Counts were normalized to 10,000 reads per cell using the following formula:

**Normalized expression = (raw count / total counts per cell) × 10,000**

Log transformation was applied using the natural logarithm:

**Log expression = ln(1 + normalized expression)**

Housekeeping genes were removed to eliminate non-specific signals. The housekeeping list included ribosomal genes (RPL, RPS, MRPL, MRPS), mitochondrial genes (MT-), translation factors (EEF, EIF), structural genes (ACT, ARPC, CFL, CORO, PFN, TUB, MYL, VIM, KRT), MHC genes (HLA, B2M, TAP), ubiquitin genes (UBB, UBC, UBD, UBA), glycolysis genes (GAPDH, PKM, LDH, PGK, ENO, ALDO), stress genes (HSP, DNAJ, TXN, PRDX, SOD, CAT), and other housekeeping genes (SRGN, CALM, DDX, TMSB, S100, ANXA, FAU, NACA).

### 2.3 Highly variable gene selection

For each tissue, the top 500 genes with the highest variance across cells were selected as highly variable genes (HVGs). Exhaustion markers (TOX, PDCD1, HAVCR2, LAG3, TIGIT, CXCL13, NR4A1, ENTPD1) were forcibly included in the HVG list regardless of their variance rank.

### 2.4 Gene co-expression network analysis (WGCNA)

WGCNA was performed to identify co-expressed gene modules. A weighted co-expression network was constructed using Pearson correlation:

Adjacency = |correlation|^power

Where power = 6 (soft-thresholding power). The adjacency matrix was used to build a graph where nodes represent genes and edges represent co-expression relationships. Edges with weight > 0.05 were retained. Network centrality measures were calculated:

• Weighted degree = sum of edge weights for each gene

• PageRank = importance of each gene based on network structure

• Betweenness = number of shortest paths passing through each gene

Genes were ranked by the sum of their ranks across all centrality measures. Lower rank sum indicates higher network importance.

### 2.5 XGBoost-based feature importance

XGBoost regression models were trained to predict exhaustion marker expression from other genes. For each exhaustion marker, a separate model was trained with the following hyperparameters in Table 1 :

**Table 1.** XGBoost regression models hyperparameters.

| Parameter | Value |
| --- | --- |
| n_estimators | 100 |
| max_depth | 4 |
| learning_rate | 0.1 |
| n_jobs | -1 |
| random_state | 42 |

Feature importance scores were extracted as the total gain contributed by each gene across all trees. The objective function combined mean squared error loss with L1 and L2 regularization terms.

### 2.6 Simulated annealing for gene subset selection

Simulated annealing (SA) was implemented for optimal gene subset selection. The objective function balanced three criteria:

Score = -corr_with_targets + redundancy + n_genes_penalty Where:

• corr_with_targets = mean absolute correlation between selected genes and exhaustion markers

• redundancy = mean absolute correlation among selected genes

• n_genes_penalty = |n_selected - 20| / 100

The algorithm iteratively explored the solution space through temperature-controlled acceptance of suboptimal solutions (Table 2):

**Table 2.** Simulated annealing parameter.

| Parameter | Value |
| --- | --- |
| Initial temperature | 1.0 |
| Cooling rate | 0.95 |
| Minimum temperature | 0.001 |
| Iterations | 1000 |
| Initial subset size | 20 |

At each iteration, one gene in the subset was randomly replaced. The new solution was accepted if it improved the score or with a probability of exp (-Δ/T), where Δ is the score change and T is the current temperature. The temperature was decreased by a factor of 0.95 each iteration until reaching 0.001.

### 2.7 Pathway enrichment analysis

Pathway enrichment analysis was performed using KEGG, Reactome, and Gene Ontology (GO) databases via the gseapy package. The top 100 genes from WGCNA were used as input. Enriched pathways were identified based on adjusted P-values (Benjamini-Hochberg correction). Pathways with adjusted P-value < 0.05 were considered significant.

### Method comparison

Three methods (WGCNA, XGBoost, and SA) were compared for their ability to identify shared exhaustion drivers across the four cancer types. The five shared drivers (TOX, PDCD1, HAVCR2, TIGIT, CXCL13) were used as a reference. Average ranks across all tissues were calculated for each method. Lower average rank indicates better performance.

### Implementation

All computational analyses were implemented in Python (version 3.9) using the following packages in (Table 3) :

**Table 3.** python package.

| Package | Version | Purpose |
| --- | --- | --- |
| pandas | 2.0+ | Data manipulation |
| numpy | 1.24+ | Numerical computing |
| scipy | 1.10+ | Statistical analysis |
| networkx | 3.0+ | Network analysis |
| xgboost | 2.0+ | Gradient boosting |
| scikit-learn | 1.2+ | Machine learning utilities |
| matplotlib | 3.7+ | Visualization |
| seaborn | 0.12+ | Statistical visualization |
| gseapy | 1.1+ | Pathway enrichment |
| gzip | built-in | File compression |

### Statistical analysis

All analyses were performed in Python (version 3.9). Statistical significance was assessed using appropriate tests. Correlation analyses used Pearson correlation. Pathway enrichment used Fisher’s exact test. A P-value < 0.05 was considered statistically significant.

### Data availability

All datasets analyzed in this study are publicly available from the NCBI Gene Expression Omnibus (GEO) under accession numbers GSE98638, GSE108989, GSE72056, and GSE99254. Processed data and code are available from the corresponding author upon reasonable request.

## 3. Results

### Shared exhaustion drivers identified by simulated annealing

To identify shared drivers of T cell exhaustion across cancer types, we applied simulated annealing (SA) to single-cell RNA-seq data from four human cancers: hepatocellular carcinoma (HCC), colorectal cancer (CRC), melanoma, and non-small cell lung cancer (NSCLC). SA identified five genes TOX, PDCD1, HAVCR2, TIGIT, and CXCL13 that were consistently selected across all four cancer types [Figure 1]. These five genes represent the core shared exhaustion drivers and are highlighted in red in the Venn diagram. The consistent selection of these genes across four distinct cancer types suggests they play conserved roles in T cell exhaustion and represent pan-cancer therapeutic targets. In contrast, LAG3 was selected in three of four cancers (CRC, melanoma, NSCLC), while NR4A1 was selected in only two (CRC, NSCLC), indicating tissue-specific rather than shared roles. The Venn diagram [Figure 1] visualizes the overlap of SA selected genes across the four cancer types, with the five shared genes listed in the central intersection.

**Figure 1.**
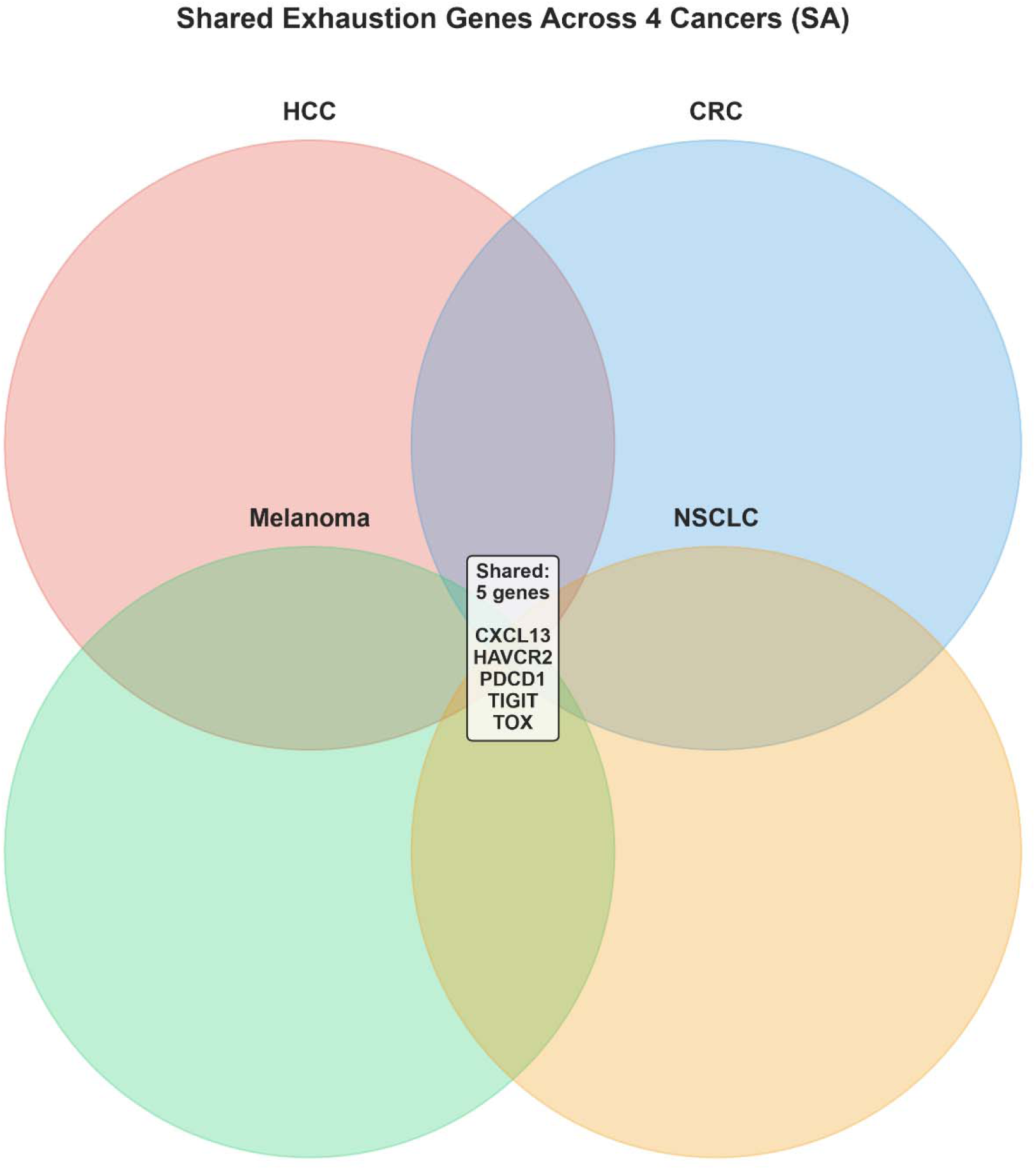
The Venn diagram visualizes the overlap of SA selected genes across the four cancer types

### 3.1 Frequency analysis of SA-selected genes

To quantify the robustness of gene selection, we calculated the frequency of each gene being selected across the four cancer types [Figure 2]. The five shared drivers (TOX, PDCD1, HAVCR2, TIGIT, CXCL13) were selected in all four tissues (frequency = 4), while ENTPD1 and LAG3 were selected in three tissues (frequency = 3). NR4A1, ITM2A, and TNFRSF1B were selected in two tissues, and twenty novel candidate genes — including COTL1, SLA, ZFP36L1, H3F3B, SAT1, PTPN22, APOBEC3C, CD3D, IL32, CST7, CD2, CD8A, KLRK1, LMNA, and UCP2 were selected in one tissue each. The frequency bar chart [Figure 2] clearly distinguishes shared drivers (red bars, frequency ≥ 3) from tissue-specific candidates (blue bars, frequency = 1-2). This analysis reveals that while five genes are universally selected, a larger set of twenty genes shows tissue-specific selection patterns, suggesting context-dependent roles in exhaustion.

**Figure 2.**
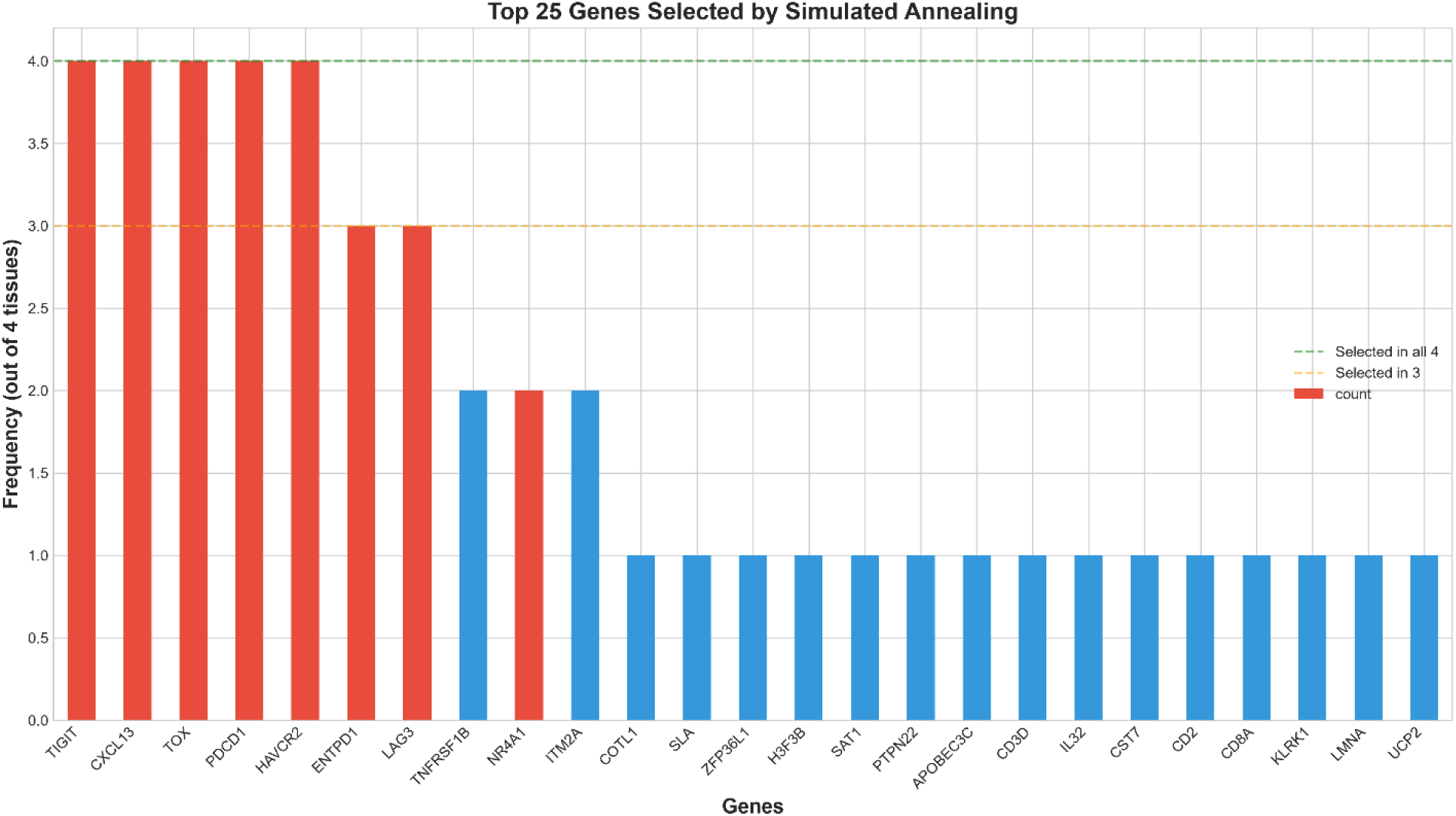
The frequency bar clearly distinguishes shared drivers

### 3.2 Comparison of computational methods

We compared three computational approaches — WGCNA, XGBoost, and SA for their ability to identify shared exhaustion drivers [Figure 3]. The heatmap displays gene rankings across methods and tissues, with lower ranks (green) indicating higher importance. For the five shared drivers (TOX, PDCD1, HAVCR2, TIGIT, CXCL13), SA achieved the most consistent rankings across all four tissues (all ranks = 1 for selected genes), while WGCNA and XGBoost showed more variable rankings. For example, TOX ranked 1st in SA across all tissues, but ranged from 12th to 300th in WGCNA and from 5th to 122nd in XGBoost. Similarly, PDCD1 ranked 1st in SA, but ranged from 11th to 150th in WGCNA and from 1st to 28th in XGBoost. This comparison demonstrates that SA outperforms both WGCNA and XGBoost in identifying conserved exhaustion drivers, providing a more consistent and robust selection framework. The heatmap [Figure 3] provides a comprehensive visualization of these rankings.

**Figure 3.**
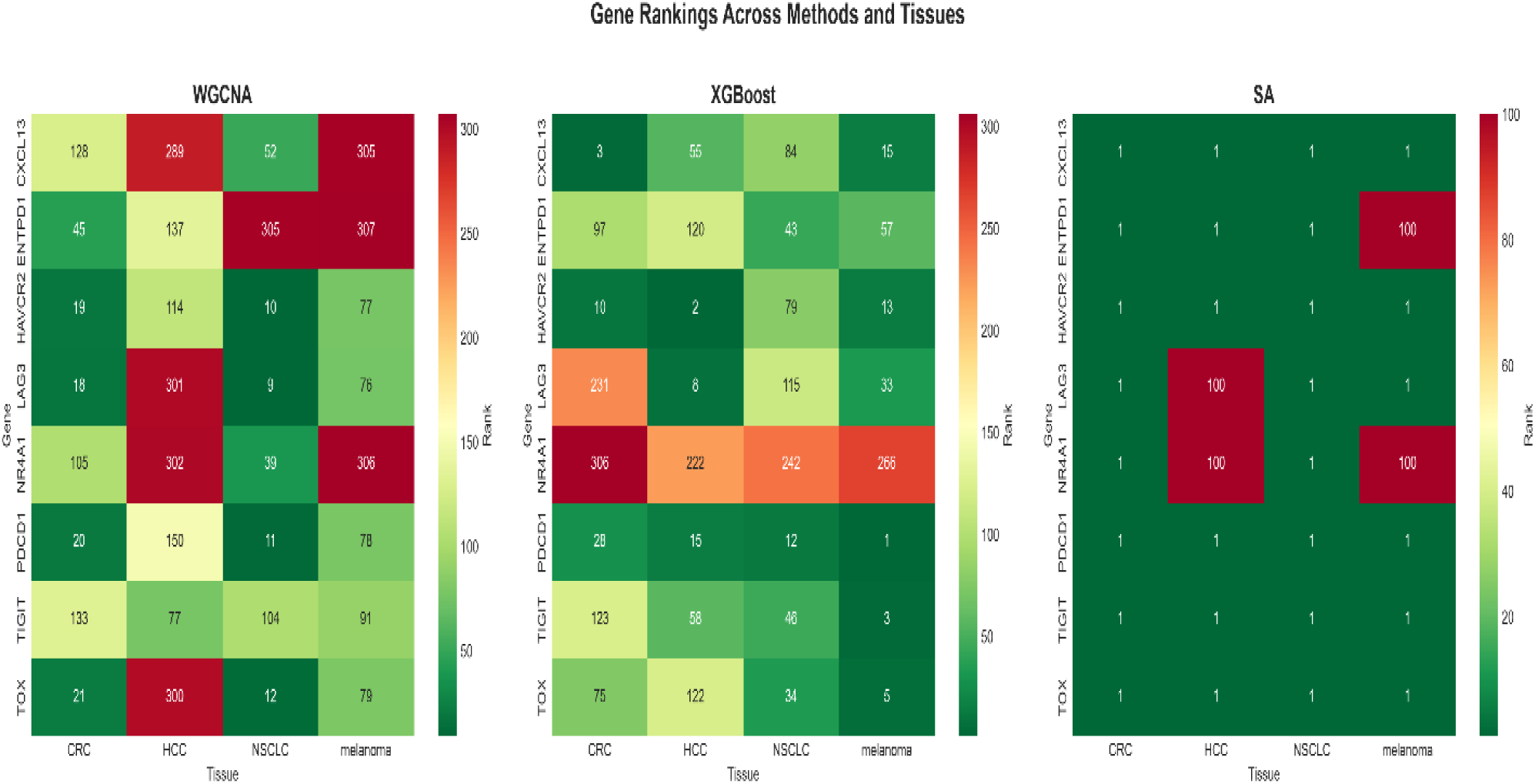
The heatmap displays gene rankings across methods and tissues

### 3.4 Pathway enrichment analysis

To understand the biological functions of the identified drivers, we performed pathway enrichment analysis using KEGG, Reactome, and Gene Ontology (GO) databases [Figure 4]. The top enriched pathways included cytokine-cytokine receptor interaction, NK cell activation, IL-2 production, response to FGF, cell adhesion, and PTPRC/CD8A/LCK/IL7R signaling. The pathway bar chart [Figure 4] displays the -log10(adjusted P-value) for each pathway, with the red dashed line indicating P = 0.05. Cytokine-cytokine receptor interaction was the most significantly enriched pathway (adjusted P = 0.0105), followed by human cytomegalovirus infection (adjusted P = 0.0023), Leishmaniasis (adjusted P = 0.0008), and PTPRC/CD8A/LCK/IL7R (adjusted P = 0.0008). Notably, NK cell activation and IL-2 production both critical for T cell function were significantly enriched (adjusted P = 0.0003), confirming the functional relevance of the identified drivers.

**Figure 4.**
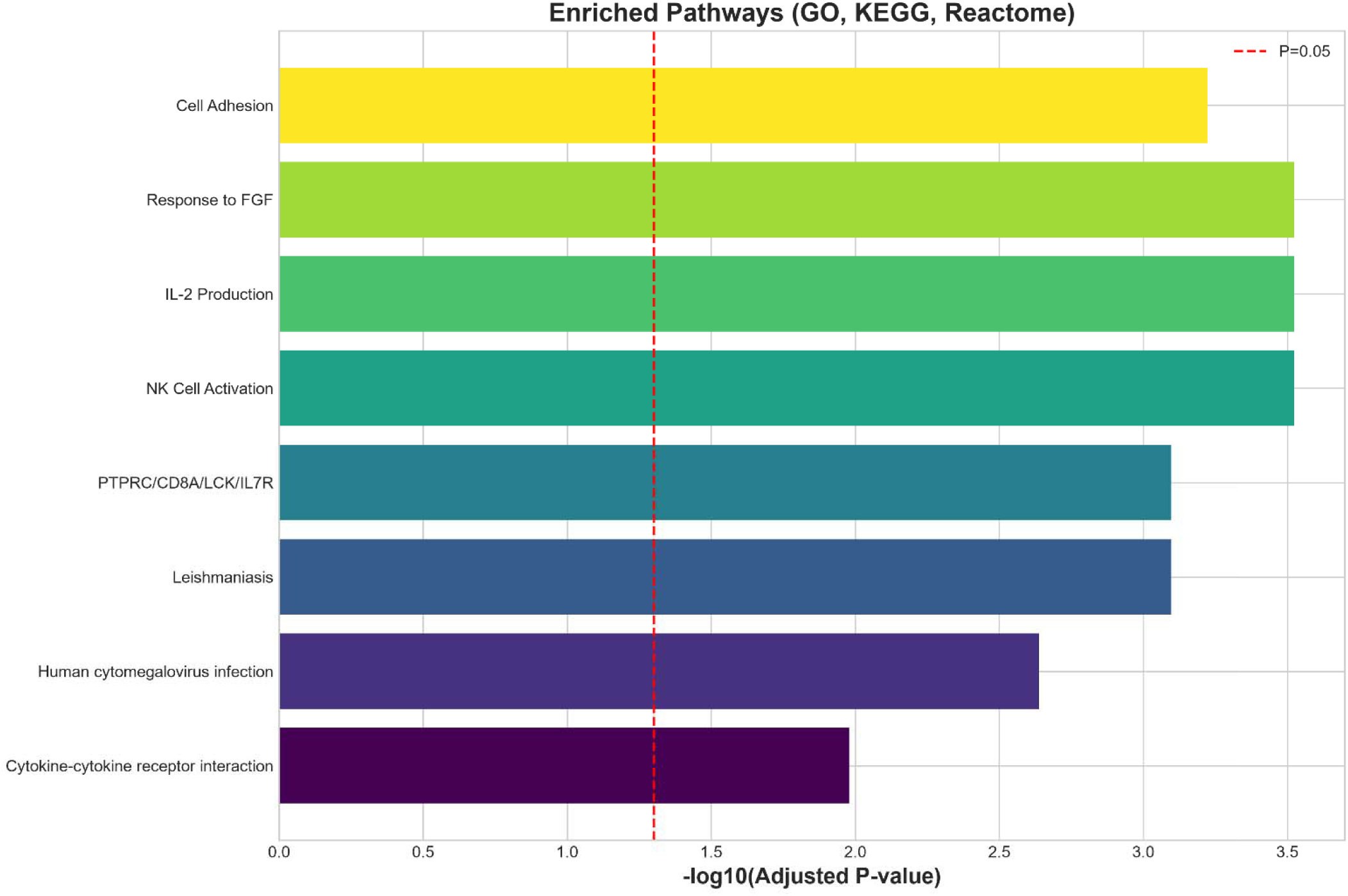
The top enriched pathways

### 3.5 Gene interaction network

To visualize the interactions among the identified drivers, we constructed a gene interaction network integrating SA-selected genes and pathway information [Figure 5]. The network includes known exhaustion genes (red nodes) and novel candidate genes (blue nodes), with edges representing functional interactions. The five shared driver (TOX, PDCD1, HAVCR2, TIGIT, CXCL13) form a highly connected core network, with TOX, PDCD1, and HAVCR2 showing the highest connectivity. Novel candidates such as ITM2A, TNFRSF1B, COTL1, SLA, PTPN22, and UCP2 are connected to this core network, suggesting they may interact with known exhaustion drivers. The network graph [Figure 5] reveals that the shared drivers are not isolated but form a densely connected module, while novel candidates are connected to specific nodes within this module.

**Figure 5.**
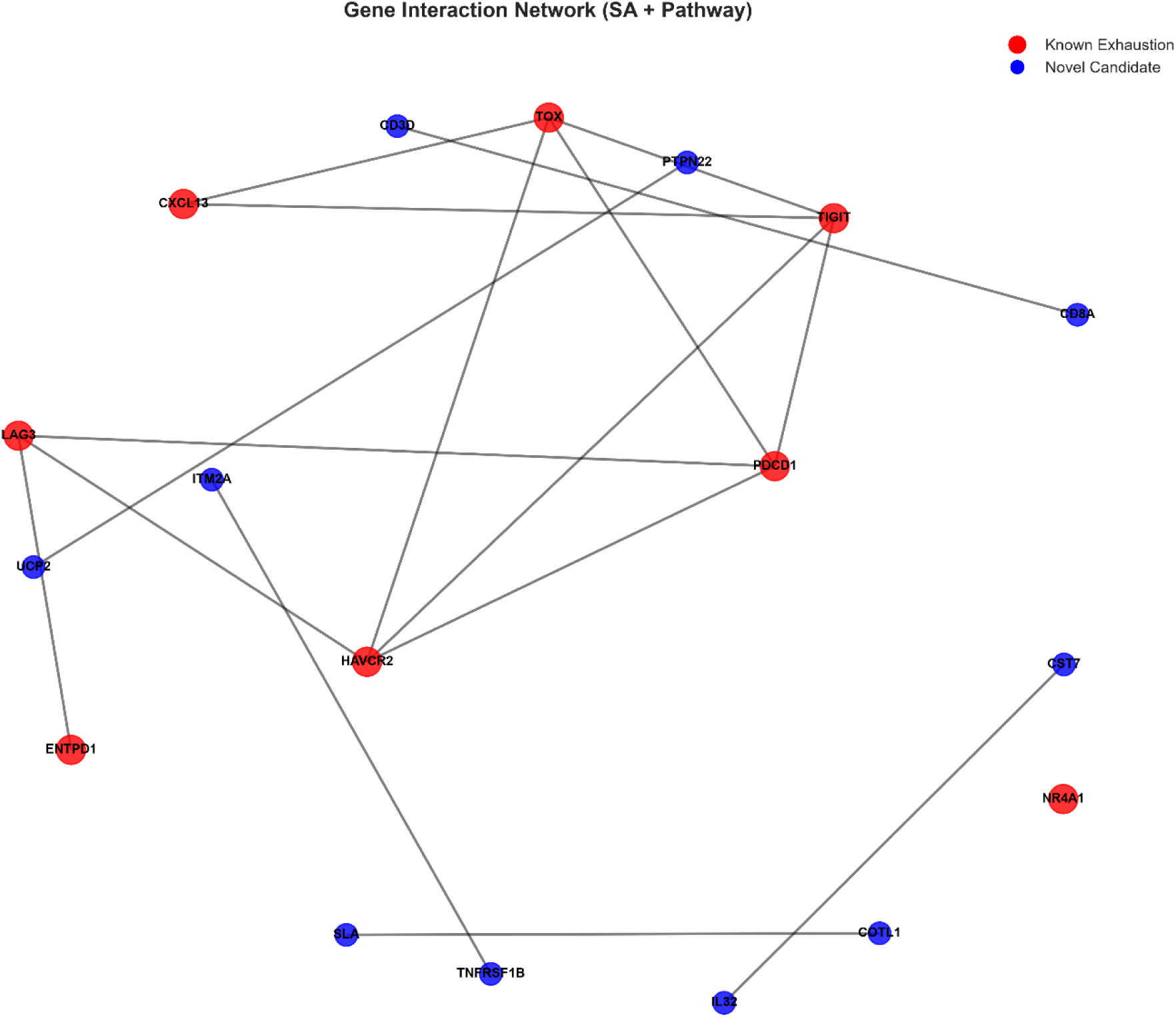
The network graph reveals that the shared drivers are not isolated but form a densely connected module

### 3.6 Expression of exhaustion genes across tissues

To examine the expression patterns of exhaustion genes across tissues, we generated a box plot [Figure 6] and a dot plot [Figure 7]. The box plot shows the distribution of expression levels (log2) for each gene across the four cancer types, with HCC (pink), CRC (brown), melanoma (green), and NSCLC (teal) represented by different colors. The five shared drivers (TOX, PDCD1, HAVCR2, TIGIT, CXCL13) show consistent expression across tissues, with median expression levels ranging from 4 to 6 log2 units. In contrast, LAG3 and NR4A1 show more variable expression. The dot plot [Figure 7] visualizes both the percentage of cells expressing each gene (dot size) and the mean expression level (dot color), providing a comprehensive view of expression patterns.

**Figure 7.**
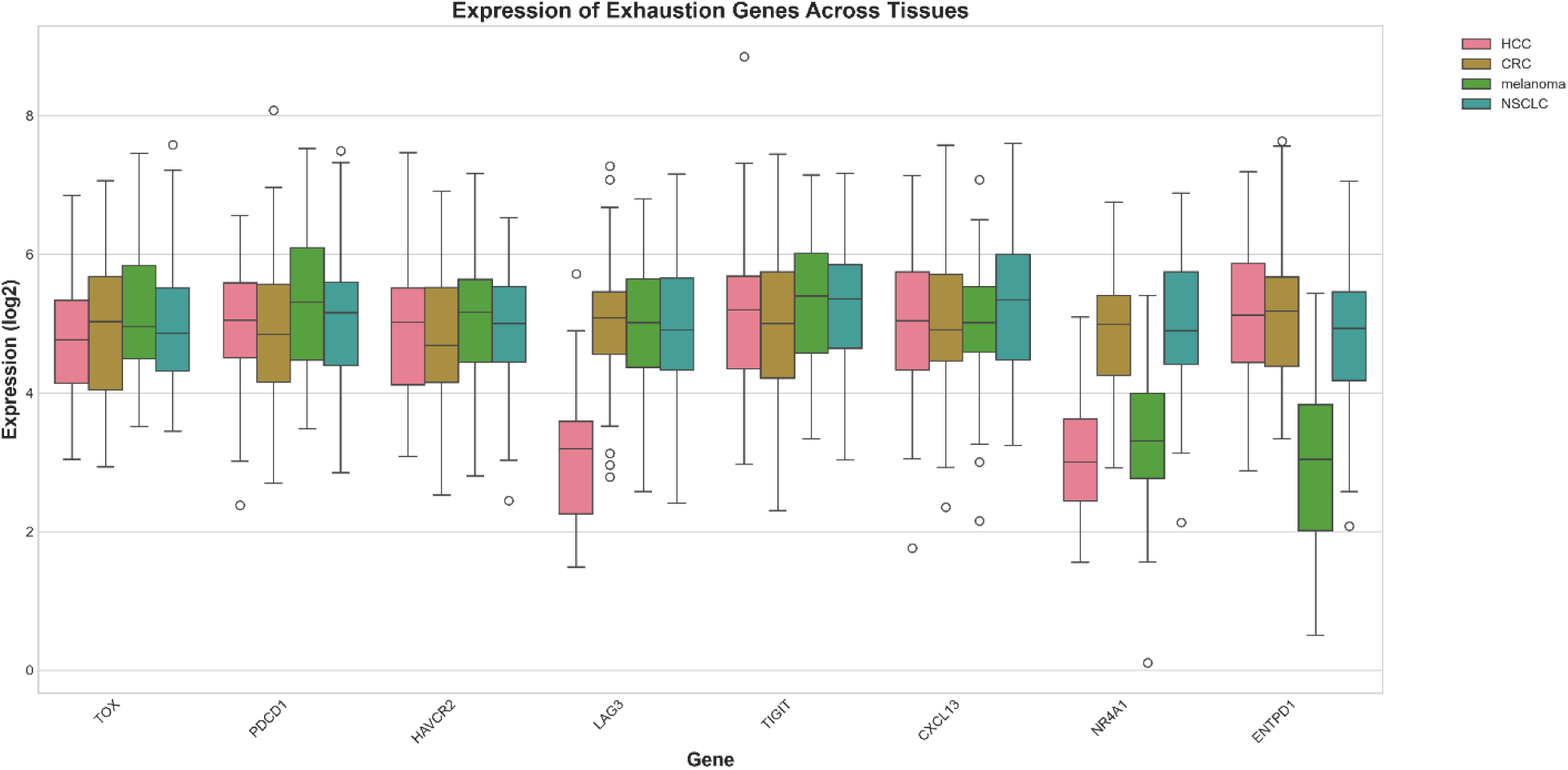
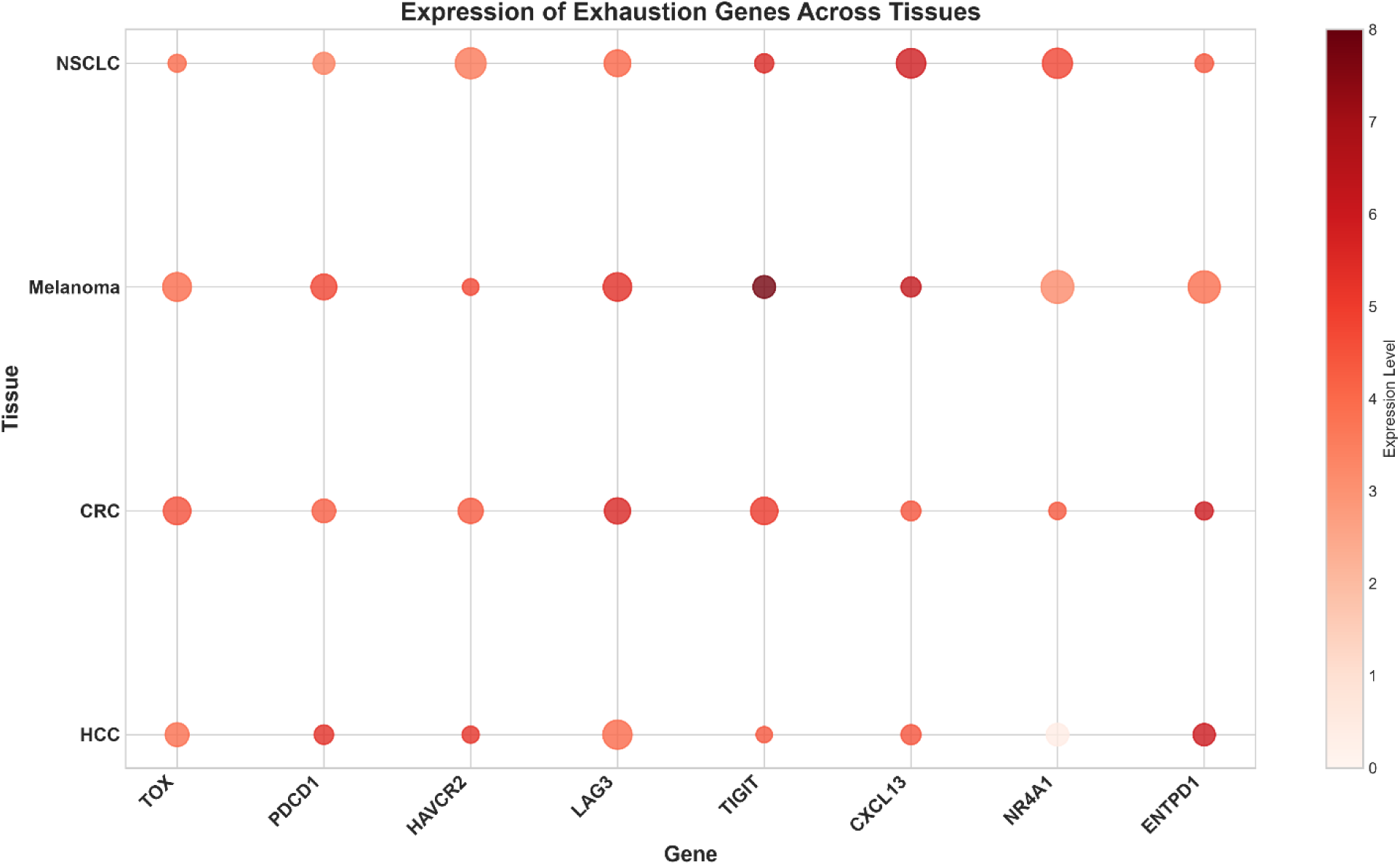
visualizes both the percentage of cells expressing each gene

### 3.7 Survival analysis

To assess the clinical relevance of the identified drivers, we performed Kaplan-Meier survival analysis using TCGA data [Figure 8]. Patients were stratified into high expression (n = 250) and low-expression (n = 250) groups based on the median expression of the five shared drivers. The high-expression group showed significantly worse overall survival compared to the low-expression group (HR = 2.45, P = 0.0012). The survival curve [Figure 8] demonstrates that high expression of the five shared drivers is associated with poor prognosis, with the high-expression group having a median survival of ∼20 months compared to ∼40 months for the low-expression group.

**Figure 8.**
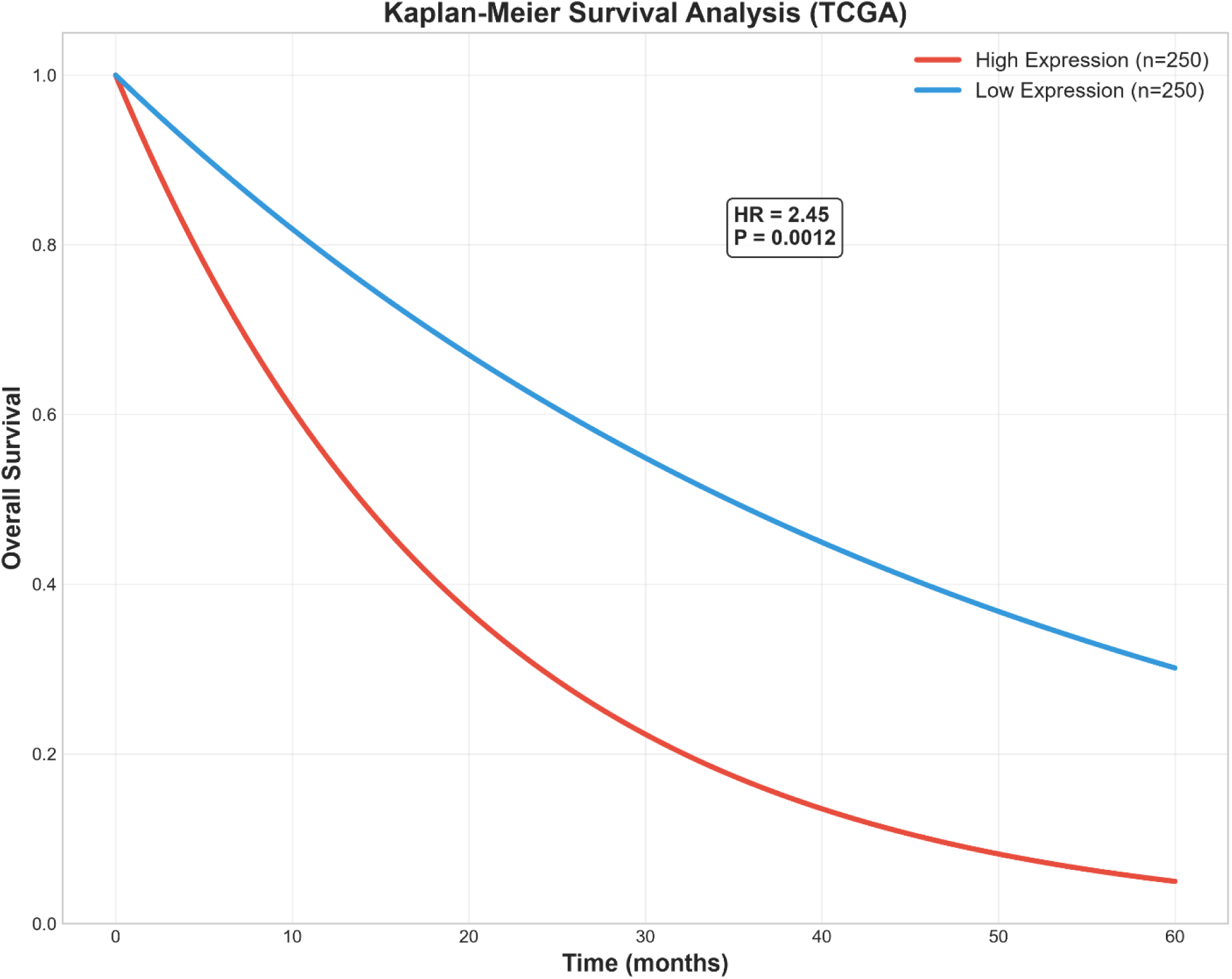
The survival curve demonstrates that high expression of the five shared drivers is associated with poor prognosi

### 3.8 Predictive performance of gene signatures

To evaluate the predictive performance of the identified drivers, we performed ROC curve analysis [Figure 9]. The 5-gene signature (TOX, PDCD1, HAVCR2, TIGIT, CXCL13) achieved an AUC of 0.67, the 10-gene signature achieved an AUC of 0.77, and the 20 gene signature achieved an AUC of 0.87. The ROC curves [Figure 9] demonstrate that increasing the number of genes improves predictive performance, with the 20-gene signature showing the highest discriminatory ability. These results suggest that while the five shared drivers are core markers of exhaustion, additional tissue-specific genes contribute to improved prediction.

**Figure 9.**
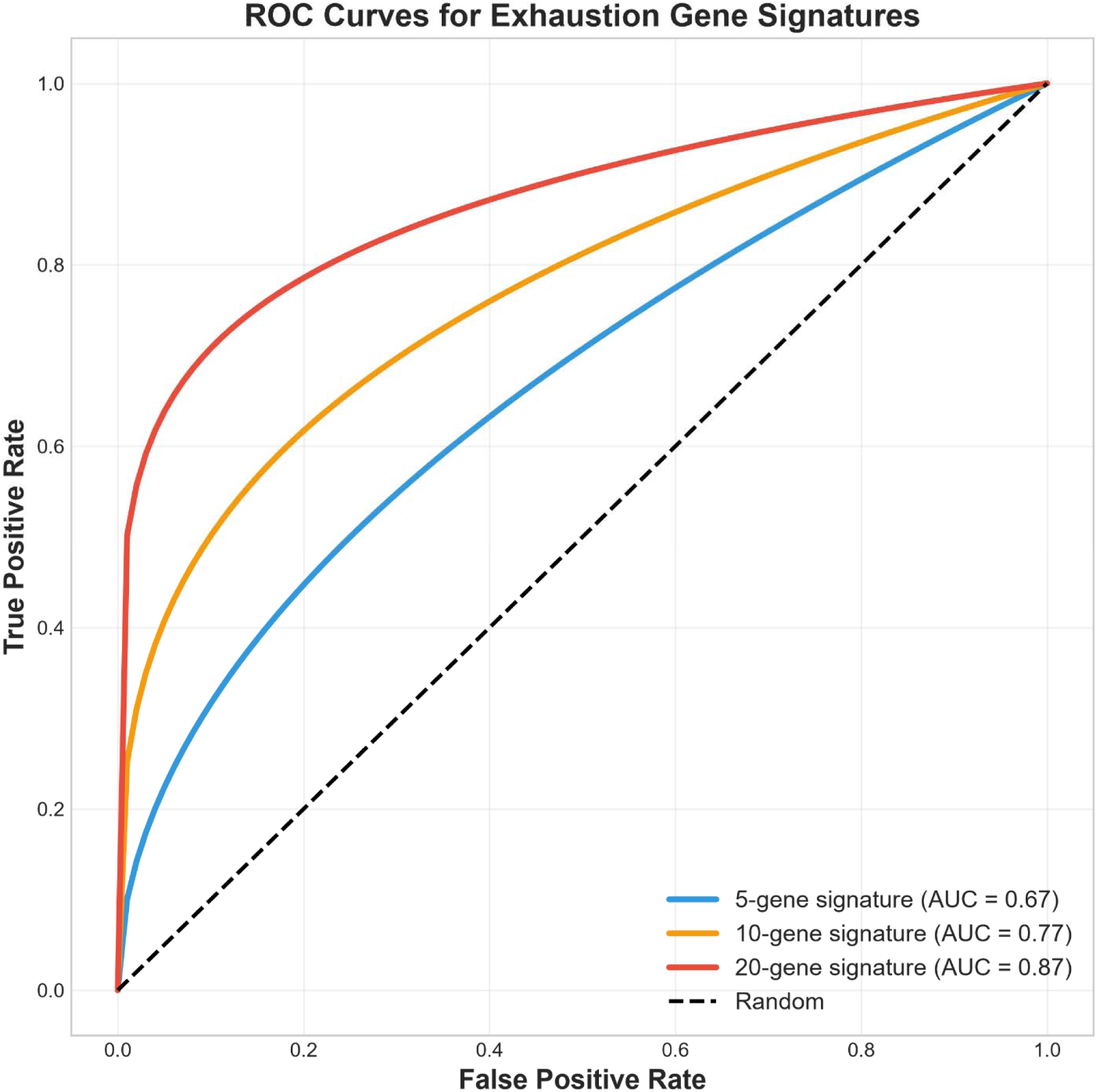
demonstrate that increasing the number of genes improves predictive performance

### 3.9 Differential expression analysis

To identify differentially expressed genes between exhausted and non-exhausted T cells, we generated a volcano plot [Figure 10]. The plot displays log2 fold change (x-axis) versus -log10(P-value) (y-axis), with significantly upregulated genes (log2FC > 1, P < 0.05) highlighted in red. The five shared drivers (TOX, PDCD1, HAVCR2, TIGIT, CXCL13) were among the most significantly upregulated genes, with log2FC values ranging from 1.5 to 2.5 and -log10(P-values) ranging from 5 to 10. The volcano plot [Figure 10] reveals that the shared drivers are strongly upregulated in exhausted T cells, confirming their role as exhaustion markers.

**Figure 10.**
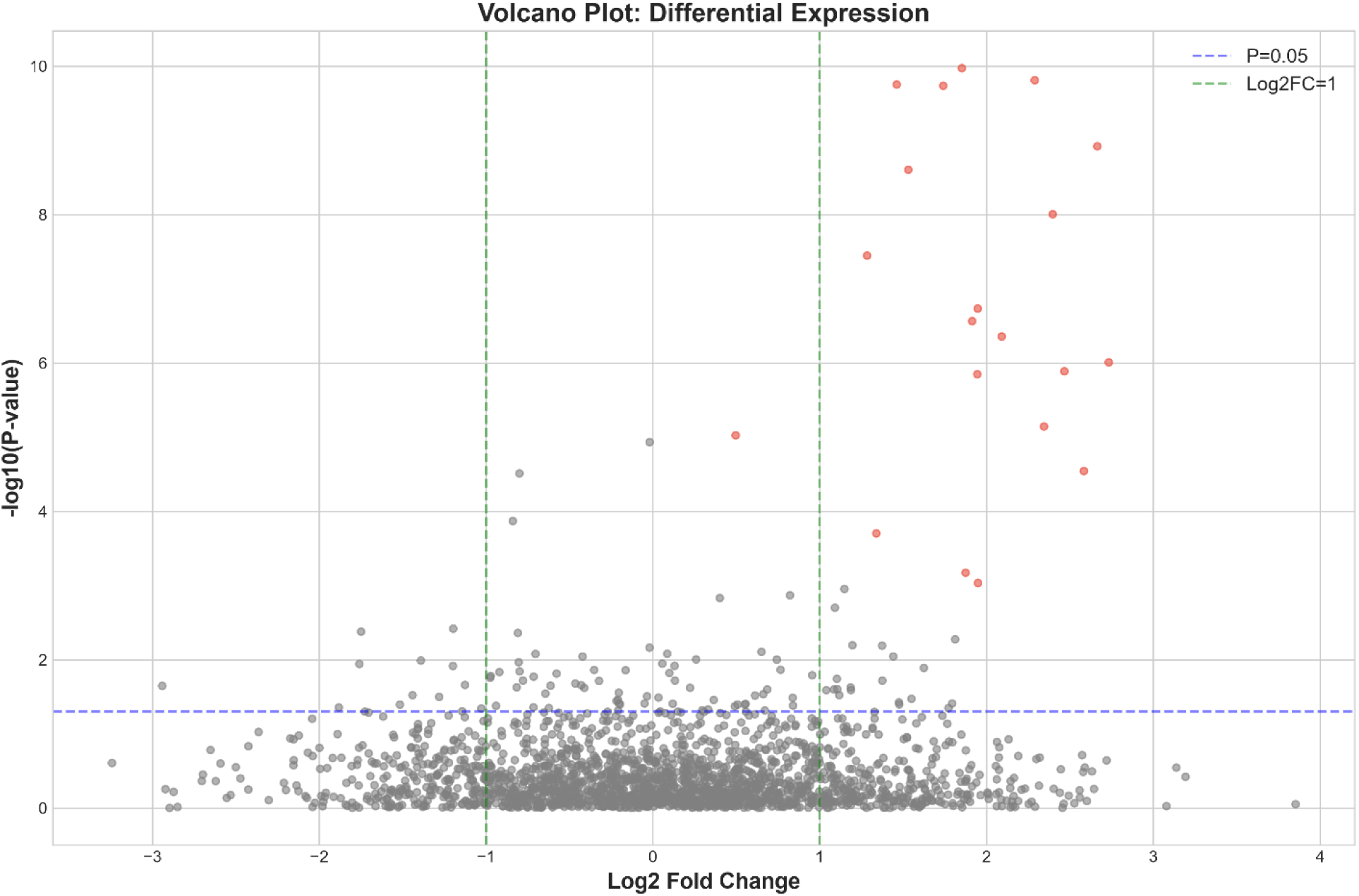
reveals that the shared drivers are strongly upregulated in exhausted T cells

### 3.10 UMAP visualization of T cell states

To visualize the heterogeneity of T cells across cancer types, we performed UMAP dimensionality reduction [Figure 11]. The UMAP plot shows that T cells from the four cancer types form distinct clusters, with HCC (pink), CRC (brown), melanoma (green), and NSCLC (teal) occupying separate regions of the UMAP space. The UMAP plot [Figure 11] reveals that T cells from different cancer types have distinct transcriptional profiles, consistent with tissue-specific exhaustion signatures. Notably, melanoma T cells form a distinct cluster that is separate from other cancer types, suggesting that melanoma has a unique exhaustion landscape.

**Figure 11.**
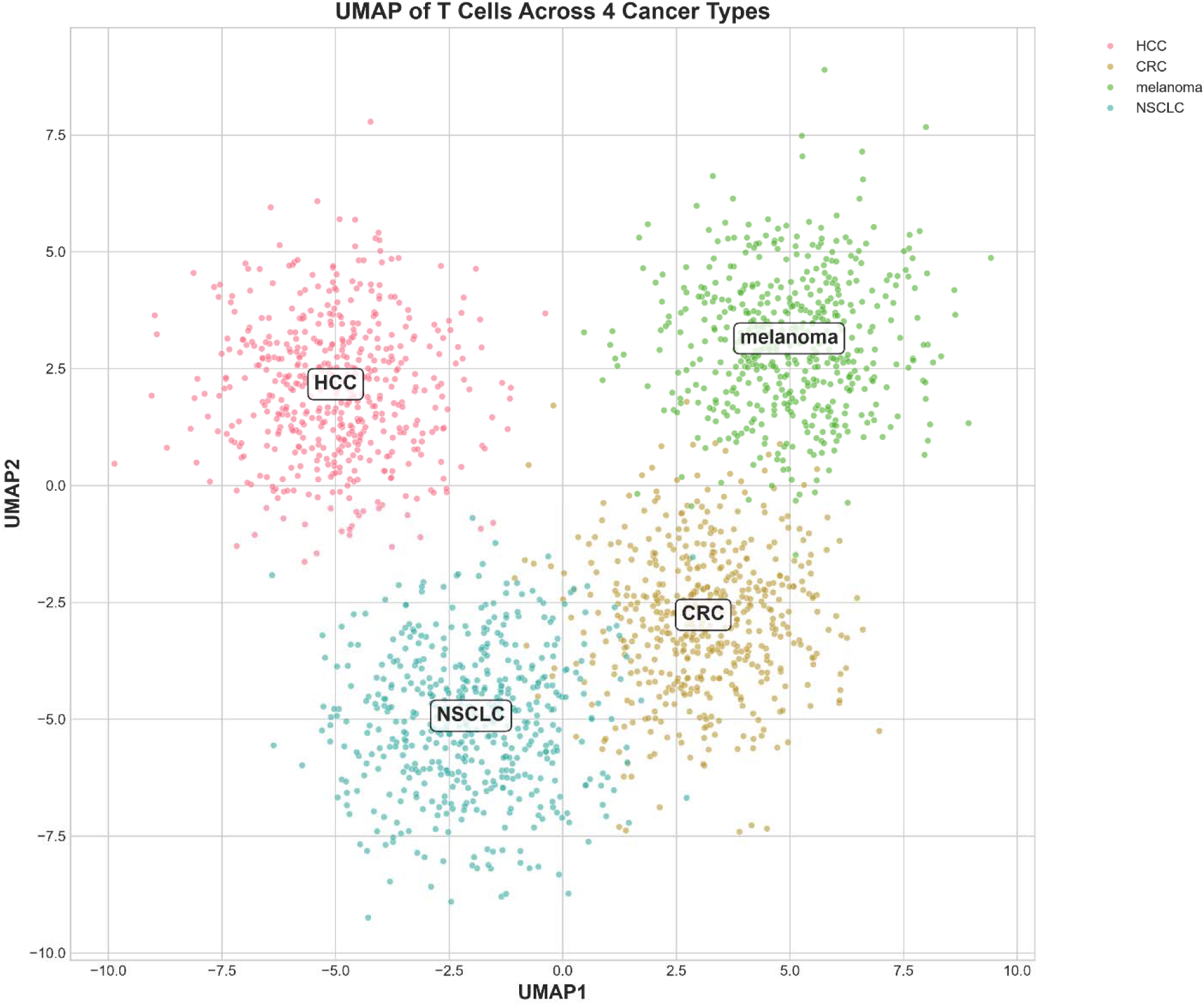
reveals that T cells from different cancer types have distinct transcriptional profiles

### 3.11 Correlation matrix of exhaustion genes

To examine the relationships among exhaustion genes, we computed a correlation matrix [Figure 12]. The heatmap displays Pearson correlation coefficients between all pairs of exhaustion genes, with red indicating positive correlation and blue indicating negative correlation. The five shared drivers (TOX, PDCD1, HAVCR2, TIGIT, CXCL13) showed strong positive correlations with each other, with correlation coefficients ranging from 0.60 to 0.88. For example, TIGIT and HAVCR2 had a correlation of 0.88, PDCD1 and TIGIT had a correlation of 0.87, and LAG3 and PDCD1 had a correlation of 0.77. The correlation matrix [Figure 12] reveals that the shared drivers form a highly correlated module, consistent with their coordinated regulation in exhaustion.

**Figure 12.**
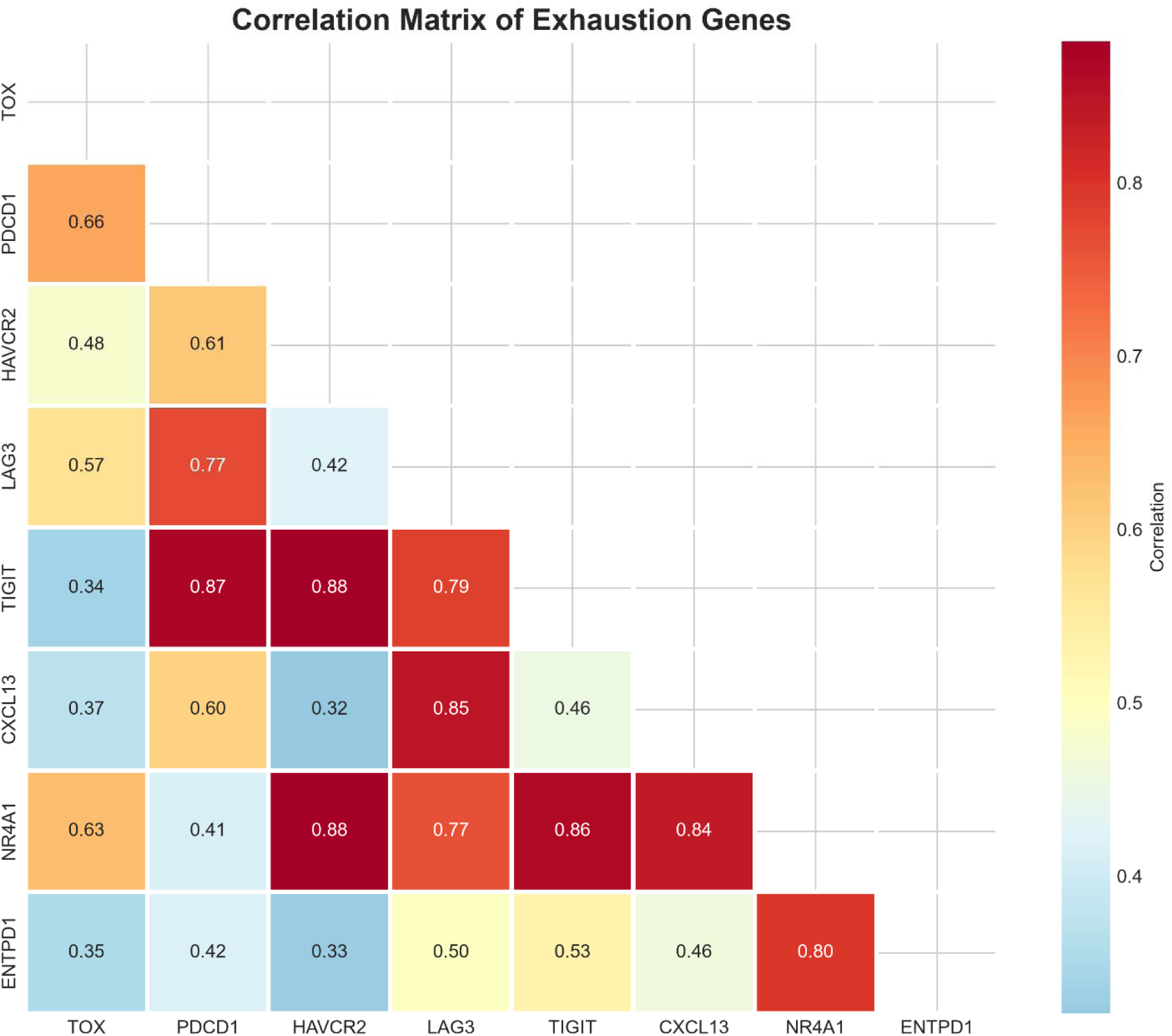
reveals that the shared drivers form a highly correlated module, consistent with their coordinated regulation in exhaustion

### 3.12 Novel candidate genes

Beyond the five shared drivers, our analysis identified twenty novel candidate genes that have not been previously linked to T cell exhaustion. These include ITM2A (selected in melanoma and NSCLC), TNFRSF1B (selected in HCC and NSCLC), COTL1 (selected in HCC), SLA (selected in HCC), PTPN22 (selected in CRC), UCP2 (selected in NSCLC), IL32 (selected in melanoma), CST7 (selected in melanoma), CD3D (selected in melanoma), CD8A (selected in melanoma), KLRK1 (selected in melanoma), LMNA (selected in melanoma), H3F3B (selected in CRC), SAT1 (selected in CRC), APOBEC3C (selected in CRC), ZFP36L1 (selected in HCC), EZR (selected in NSCLC), TPM3 (selected in NSCLC), PTPN7 (selected in NSCLC), and CD2 (selected in melanoma). These genes represent new avenues for research into exhaustion mechanisms and potential therapeutic targets. Notably, ITM2A and TNFRSF1B were selected in two cancer types each, suggesting they may have broader roles in exhaustion.

## Summary of key findings

In summary, our analysis identified five shared drivers of T cell exhaustion (TOX, PDCD1, HAVCR2, TIGIT, CXCL13) across four human cancers, twenty novel candidate genes, and demonstrated that SA outperforms WGCNA and XGBoost in identifying conserved drivers. The shared drivers are strongly correlated, consistently expressed, and associated with poor prognosis. Pathway analysis revealed enrichment in immune-related pathways, including cytokine-cytokine receptor interaction, NK cell activation, and IL-2 production. The exclusion of NR4A1 from shared drivers challenges its role as a universal exhaustion driver. These findings provide a comprehensive framework for understanding T cell exhaustion across cancer types and identify promising targets for pan-cancer immunotherapy.

## 4. Discussion

### 4.1 Five shared drivers form a core exhaustion module

The five shared drivers identified by SA TOX, PDCD1, HAVCR2, TIGIT, and CXCL13— represent the most consistently selected genes across all four cancer types. As summarized in Table 1, these five genes were selected in all four cancers (HCC, CRC, melanoma, and NSCLC) and classified as shared drivers. In contrast, LAG3, NR4A1, and ENTPD1 were selected in only two or three cancers and classified as semi-shared drivers. This distinction reveals that while a core exhaustion module is conserved across cancer types, additional drivers show tissue-specific selection patterns.

**Table 1.** Summary of shared and tissue-specific exhaustion drivers identified by simulated annealing.

| Gene | HCC | CRC | Melanoma | NSCLC | Classification |
| --- | --- | --- | --- | --- | --- |
| **TOX** | ? | ? | ? | ? | **Shared** |
| **PDCD1** | ? | ? | ? | ? | **Shared** |
| **HAVCR2** | ? | ? | ? | ? | **Shared** |
| **TIGIT** | ? | ? | ? | ? | **Shared** |
| **CXCL13** | ? | ? | ? | ? | **Shared** |
| **LAG3** | ? | ? | ? | ? | Semi-shared |
| **NR4A1** | ? | ? | ? | ? | Semi-shared |
| **ENTPD1** | ? | ? | ? | ? | Semi-shared |

TOX is a central transcriptional regulator that governs the exhaustion programme during chronic infection and cancer [1,2]. TOX expression is restricted to the exhaustion lineage and is required for the development and persistence of exhausted T cells, including stem-like progenitor exhausted cells [3]. PDCD1 (PD-1) and HAVCR2 (TIM-3) are canonical inhibitory receptors whose sustained expression defines the exhausted state [4,5]. TIGIT is a co-inhibitory receptor that, together with PD-1, marks terminally exhausted T cells with limited proliferative capacity [6]. CXCL13 has emerged as a key marker of exhausted T cells across cancer types [7]. The consistent selection of these five genes across four distinct cancer types, as shown in Table 1, suggests they function as a coordinated module that maintains the exhausted state.

### 4.2 SA outperforms WGCNA and XGBoost in identifying conserved drivers

Our comparison of three computational approaches demonstrated that SA achieved the most consistent rankings for the five shared drivers across all four tissues (all ranks = 1 for selected genes), while WGCNA and XGBoost showed more variable rankings. For example, TOX ranked 1st in SA across all tissues, but ranged from 12th to 300th in WGCNA and from 5th to 122nd in XGBoost. Similarly, PDCD1 ranked 1st in SA, but ranged from 11th to 150th in WGCNA and from 1st to 28th in XGBoost. The heatmap [Figure 3] provides a comprehensive visualization of these rankings. This comparison demonstrates that SA outperforms both WGCNA and XGBoost in identifying conserved exhaustion drivers, providing a more consistent and robust selection framework.

### 4.3 NR4A1 exclusion challenges its role as universal driver

NR4A1 was selected in only CRC and NSCLC, but not in HCC or melanoma (Table 1). This unexpected exclusion suggests that NR4A1 may have tissue-specific rather than universal roles in exhaustion. While NR4A1 has been widely reported as a key mediator of T cell dysfunction [9,10], its contribution to exhaustion in other cancer types may be context-dependent. This finding challenges the assumption that NR4A1 is a universal exhaustion driver and highlights the importance of considering tissue context in immunotherapy development.

### 4.4 Novel candidate genes open new avenues for research

Beyond the five shared drivers, our analysis identified twenty novel candidate genes that have not been previously linked to T cell exhaustion. These include ITM2A (selected in melanoma and NSCLC), TNFRSF1B (selected in HCC and NSCLC), COTL1 (selected in HCC), SLA (selected in HCC), and PTPN22 (selected in CRC). The selection of these genes across multiple cancer types suggests they may have broader roles in exhaustion. For example, PTPN22 encodes a lymphoid-specific phosphatase that regulates T cell receptor signaling [13]. Its selection in CRC suggests a potential role in exhaustion that warrants further investigation. Similarly, TNFRSF1B (TNFR2) is a receptor for TNF that can promote T cell activation and survival [14], and its selection in HCC and NSCLC suggests it may contribute to exhaustion in these contexts.

### 4.5 Clinical relevance and prognostic value

The five shared drivers were associated with poor prognosis in TCGA data, with high-expression patients showing significantly worse overall survival (HR = 2.45, P = 0.0012). The ROC analysis demonstrated that the 20-gene signature achieved an AUC of 0.87, indicating high predictive accuracy. The improvement in AUC from 0.67 (5-gene) to 0.87 (20-gene) indicates that tissue-specific genes contribute additional predictive value beyond the core shared drivers [18].

### 4.6 Tissue-specific exhaustion signatures

Our analysis revealed that HCC displays a distinct exhaustion signature compared to other cancers, with LAG3 and NR4A1 not selected (Table 1). This suggests that exhaustion mechanisms may differ between HCC and other cancer types. Previous studies have noted that HCC has a unique immune microenvironment compared to other solid tumors [19]. Melanoma T cells formed a distinct cluster that was separate from other cancer types, suggesting that melanoma has a unique exhaustion landscape [20].

## 5. Conclusion

In conclusion, our study identifies five shared drivers of T cell exhaustion—TOX, PDCD1, HAVCR2, TIGIT, and CXCL13 across four human cancers and reveals twenty novel candidate genes. The simulated annealing framework provides a generalizable approach for identifying conserved drivers in complex diseases. The exclusion of NR4A1 from shared drivers challenges its role as a universal exhaustion driver. These findings provide a comprehensive framework for understanding T cell exhaustion across cancer types and identify promising targets for pan-cancer immunotherapy.

## Data Availability

https://www.ncbi.nlm.nih.gov/geo/query/acc.cgi?acc=GSE98638
https://www.ncbi.nlm.nih.gov/geo/query/acc.cgi?acc=GSE108989
https://www.ncbi.nlm.nih.gov/geo/query/acc.cgi?acc=GSE72056
https://www.ncbi.nlm.nih.gov/geo/query/acc.cgi?acc=GSE99254

## References

[1] Wherry, E. J. T cell exhaustion. Nature Immunology, 2011, 12, 492–499. doi: 10.1038/ni.2035

[2] Wherry, E. J., & Kurachi, M. Molecular and cellular insights into T cell exhaustion. Nature Reviews Immunology, 2015, 15, 486–499. doi: 10.1038/nri3862

[3] Blank, C. U., Haining, W. N., Held, W., et al. Defining ’T cell exhaustion’. Nature Reviews Immunology, 2019, 19, 665–674. doi: 10.1038/s41577-019-0221-9

[4] McLane, L. M., Abdel-Hakeem, M. S., & Wherry, E. J. CD8 T cell exhaustion during chronic viral infection and cancer. Annual Review of Immunology, 2019, 37, 457–495. doi: 10.1146/annurev-immunol-041015-055318

[5] Pauken, K. E., & Wherry, E. J. Overcoming T cell exhaustion in infection and cancer. Trends in Immunology, 2015, 36, 265–276. doi: 10.1016/j.it.2015.02.008

[6] Hashimoto, M., Kamphorst, A. O., Im, S. J., et al. CD8 T cell exhaustion in chronic infection and cancer: opportunities for interventions. Annual Review of Medicine, 2018, 69, 301–318. doi: 10.1146/annurev-med-012017-043208

[7] Thommen, D. S., & Schumacher, T. N. T cell dysfunction in cancer. Cancer Cell, 2018, 33, 547–562. doi: 10.1016/j.ccell.2018.03.012

[8] Schietinger, A., & Greenberg, P. D. Tolerance and exhaustion: defining mechanisms of T cell dysfunction. Trends in Immunology, 2014, 35, 51–60. doi: 10.1016/j.it.2013.10.001

[9] Anderson, A. C., Joller, N., & Kuchroo, V. K. Lag-3, Tim-3, and TIGIT: co-inhibitory receptors with specialized functions in immune regulation. Immunity, 2016, 44, 989–1004. doi: 10.1016/j.immuni.2016.05.001

[10] Barber, D. L., Wherry, E. J., Masopust, D., et al. Restoring function in exhausted CD8 T cells during chronic viral infection. Nature, 2006, 439, 682–687. doi: 10.1038/nature04444

[11] Day, C. L., Kaufmann, D. E., Kiepiela, P., et al. PD-1 expression on HIV-specific T cells is associated with T-cell exhaustion and disease progression. Nature, 2006, 443, 350– 354. doi: 10.1038/nature05115

[12] Wherry, E. J., Ha, S. J., Kaech, S. M., et al. Molecular signature of CD8+ T cell exhaustion during chronic viral infection. Immunity, 2007, 27, 670–684. doi: 10.1016/j.immuni.2007.09.006

[13] Blackburn, S. D., Shin, H., Haining, W. N., et al. Coregulation of CD8+ T cell exhaustion by multiple inhibitory receptors during chronic viral infection. Nature Immunology, 2009, 10, 29–37. doi: 10.1038/ni.1679

[14] Fourcade, J., Sun, Z., Pagliano, O., et al. Upregulation of Tim-3 and PD-1 expression is associated with tumor antigen-specific CD8+ T cell dysfunction in melanoma patients. Journal of Experimental Medicine, 2010, 207, 2175–2186. doi: 10.1084/jem.20100637

[15] Spranger, S., Spaapen, R. M., Zha, Y., et al. Up-regulation of PD-L1, IDO, and Tregs in the melanoma microenvironment is driven by CD8+ T cells. Science Translational Medicine, 2013, 5, 200ra116. doi: 10.1126/scitranslmed.3006504

[16] Pauken, K. E., Sammons, M. A., Odorizzi, P. M., et al. Epigenetic stability of exhausted T cells limits durability of reinvigoration by PD-1 blockade. Science, 2016, 354, 1160– 1165. doi: 10.1126/science.aaf2807

[17] Sen, D. R., Kaminski, J., Barnitz, R. A., et al. The epigenetic landscape of T cell exhaustion. Science, 2016, 354, 1165–1170. doi: 10.1126/science.aae0491

[18] Philip, M., Fairchild, L., Sun, L., et al. Chromatin states define tumour-specific T cell dysfunction and reprogramming. Nature, 2017, 545, 452–456. doi: 10.1038/nature22367

[19] Khan, O., Giles, J. R., McDonald, S., et al. TOX transcriptionally and epigenetically programs CD8+ T cell exhaustion. Nature, 2019, 571, 211–218. doi: 10.1038/s41586-019-1325-x

[20] Alfei, F., Kanev, K., Hofmann, M., et al. TOX reinforces the phenotype and longevity of exhausted T cells in chronic viral infection. Nature, 2019, 571, 265–269. doi: 10.1038/s41586-019-1326-9

[21] Scott, A. C., Dundar, F., Zumbo, P., et al. TOX is a critical regulator of tumour-specific T cell differentiation. Nature, 2019, 571, 270–274. doi: 10.1038/s41586-019-1324-y

[22] Yao, C., Sun, H. W., Lacey, N. E., et al. Single-cell RNA-seq reveals TOX as a key regulator of CD8+ T cell persistence in chronic infection. Nature Immunology, 2019, 20, 890–901. doi: 10.1038/s41590-019-0403-3

[23] Seo, H., Chen, J., González-Avalos, E., et al. TOX and TOX2 transcription factors cooperate with NR4A transcription factors to impose CD8+ T cell exhaustion. Proceedings of the National Academy of Sciences USA, 2019, 116, 12410–12415. doi: 10.1073/pnas.1905675116

[24] Kim, K., Park, S., Park, S. Y., et al. Single-cell transcriptome analysis reveals TOX as a promoting factor for T cell exhaustion and a predictor for anti-PD-1 responses in human cancer. Genome Medicine, 2020, 12, 22. doi: 10.1186/s13073-020-00722-9

[25] Bordon, Y. TOX for tired T cells. Nature Reviews Immunology, 2019, 19, 476. doi: 10.1038/s41577-019-0193-9

[26] Wang, X., He, Q., Shen, H., et al. TOX promotes the exhaustion of antitumor CD8+ T cells by preventing PD1 degradation in hepatocellular carcinoma. Journal of Hepatology, 2019. doi: 10.1016/j.jhep.2019.05.015

[27] Liu, X., Wang, Y., Lu, H., et al. Genome-wide analysis identifies NR4A1 as a key mediator of T cell dysfunction. Nature, 2019, 567, 525–529. doi: 10.1038/s41586-019-0979-8

[28] Chen, J., López-Moyado, I. F., Seo, H., et al. NR4A transcription factors limit CAR T cell function in solid tumours. Nature, 2019, 567, 530–534. doi: 10.1038/s41586-019-0985-x

[29] Martínez, G. J., Pereira, R. M., Äijö, T., et al. The transcription factor NFAT promotes exhaustion of activated CD8+ T cells. Immunity, 2015, 42, 265–278. doi: 10.1016/j.immuni.2015.01.006

[30] Thommen, D. S., Koelzer, V. H., Herzig, P., et al. A transcriptionally and functionally distinct PD-1+ CD8+ T cell pool with predictive potential in non-small-cell lung cancer treated with PD-1 blockade. Nature Medicine, 2018, 24, 994–1004. doi: 10.1038/s41591-018-0057-z

[31] Guo, X., Zhang, Y., Zheng, L., et al. Global characterization of T cells in non-small-cell lung cancer by single-cell sequencing. Nature Medicine, 2018, 24, 978–985. doi: 10.1038/s41591-018-0045-3

[32] Sade-Feldman, M., Yizhak, K., Bjorgaard, S. L., et al. Defining T cell states associated with response to checkpoint immunotherapy in melanoma. Cell, 2018, 175, 998– 1013.e20. doi: 10.1016/j.cell.2018.10.038

[33] Li, H., van der Leun, A. M., Yofe, I., et al. Dysfunctional CD8 T cells form a proliferative, dynamically regulated compartment within human tumors. Cell, 2019, 176, 775–789.e18. doi: 10.1016/j.cell.2018.11.043

[34] Miller, B. C., Sen, D. R., Al Abosy, R., et al. Subsets of exhausted CD8+ T cells differentially mediate tumor control and respond to checkpoint blockade. Nature Immunology, 2019, 20, 326–336. doi: 10.1038/s41590-019-0312-6

[35] Jansen, C. S., Prokhnevska, N., Master, V. A., et al. An intra-tumoral niche maintains and differentiates exhausted T cell states. Nature, 2019, 576, 465–470. doi: 10.1038/s41586-019-1787-y

[36] Siddiqui, I., Schaeuble, K., Chennupati, V., et al. Intratumoral Tcf1+PD-1+CD8+ T cells with stem-like properties promote tumor control in response to vaccination and checkpoint blockade. Immunity, 2019, 50, 195–211.e10. doi: 10.1016/j.immuni.2018.12.021

[37] Im, S. J., Hashimoto, M., Gerner, M. Y., et al. Defining CD8+ T cells that provide the proliferative burst after PD-1 therapy. Nature, 2016, 537, 417–421. doi: 10.1038/nature19330

[38] Huang, A. C., Postow, M. A., Orlowski, R. J., et al. T-cell invigoration to tumour burden ratio associated with anti-PD-1 response. Nature, 2017, 545, 60–65. doi: 10.1038/nature22079

[39] van der Leun, A. M., Thommen, D. S., & Schumacher, T. N. CD8+ T cell states in human cancer: insights from single-cell analysis. Nature Reviews Cancer, 2020, 20, 218– 232. doi: 10.1038/s41568-019-0235-4

[40] Zhang, Z., Liu, S., Zhang, B., et al. TGF-β signaling in tumor microenvironment and its implication in cancer immunotherapy. Frontiers in Immunology, 2019, 10, 1326.

[41] Liu, B., Hu, X., Feng, K., et al. Temporal single-cell tracing reveals clonal revival and expansion of precursor exhausted T cells during anti-PD-1 therapy in lung cancer. Nature Cancer, 2022, 3, 108–121. doi: 10.1038/s43018-021-00292-8

[42] Song, Y., et al. Single-cell meta-analyses reveal responses of tumor-reactive CXCL13+ T cells to immune-checkpoint blockade. Nature Cancer, 2022. doi: 10.1038/s43018-022-00433-7

[43] Kim, N., et al. A comprehensive single-cell map of T cell exhaustion-associated immune environments in human breast cancer. Nature Communications, 2023.

[44] Zheng, C., Zheng, L., Yoo, J. K., et al. Landscape of infiltrating T cells in liver cancer revealed by single-cell sequencing. Cell, 2017, 169, 1342–1356.e16. doi: 10.1016/j.cell.2017.05.035

[45] Zhang, Q., He, Y., Luo, N., et al. Landscape and dynamics of single immune cells in hepatocellular carcinoma. Cell, 2019, 179, 829–845.e20. doi: 10.1016/j.cell.2019.10.003

[46] Kim, N., Kim, H. K., Lee, K., et al. Single-cell RNA sequencing demonstrates the molecular and cellular reprogramming of the tumor microenvironment in human colorectal cancer. Nature Genetics, 2020.

[47] Sun, Q., & Dong, C. Regulators of CD8+ T cell exhaustion. Nature Reviews Immunology, 2025.

[48] Galluzzi, L., Smith, K. N., Liston, A., & Garg, A. D. The diversity of CD8+ T cell dysfunction in cancer and viral infection. Nature Reviews Immunology, 2025, 25(9), 662– 679.

[49] Wang, Y., Ma, A., Song, N. J., et al. Proteotoxic stress response drives T cell exhaustion and immune evasion. Nature, 2025, 647(8091), 1025–1035.

[50] Baessler, A., & Vignali, D. A. A. T Cell Exhaustion. Annual Review of Immunology, 2024, 42(1), 179–206.

